# The effectiveness of digital delivery versus group-based face-to-face delivery of the English National Health Service Diabetes Prevention Programme: a non-inferiority retrospective cohort comparison study

**DOI:** 10.1101/2023.02.21.23286221

**Authors:** Antonia M. Marsden, Mark Hann, Emma Barron, Ben McGough, Elizabeth Murray, Jonathan Valabhji, Sarah Cotterill

## Abstract

**Introduction:** Face-to-face group-based diabetes prevention programmes have been shown to be effective in many settings. Digital delivery may suit some patients, but research comparing the effectiveness of digital with face-to-face delivery is scarce. The aim was to assess if digital delivery of the English National Health Service Diabetes Prevention Programme (NHS DPP) is non-inferior to group-based face-to-face delivery in terms of weight change, and evaluate factors associated with differential change. The study included those recruited to the NHS DPP in 2017-2018.

**Research design and methods:** Individual-level data from a face-to-face cohort was compared to two cohorts on a digital pilot who (i) were offered no choice of delivery mode, or (ii) chose digital over face-to-face. Changes in weight at 6 and 12 months were analysed using mixed effects linear regression, having matched participants from the digital pilot to similar participants from face-to-face.

**Results:** Weight change on the digital pilot was non-inferior to face-to-face at both time points: it was similar in the comparison of those with no choice (difference in weight change: −0.284kg [95% CI: −0.712, 0.144] at 6 months) and greater in digital when participants were offered a choice (−1.165kg [95% CI: −1.841, −0.489]). Interactions between delivery mode and sex, ethnicity, age and deprivation were observed.

**Conclusions:** Digital delivery of the NHS DPP achieved weight loss at least as good as face-to-face. Patients who were offered a choice and opted for digital experienced better weight loss, compared to patients offered face-to-face only.

## INTRODUCTION

Diabetes prevention is a major public health objective as the prevalence of type 2 diabetes (T2D) is increasing globally.^1-3^ Diabetes prevention programmes (DPPs), offering behaviour change support with weight loss, dietary change and increased exercise, can be effective in reducing T2D onset in those at high risk,^4, 5^ but most of the evidence to date is based on face-to-face delivery, often in small groups.

Group delivery is disliked by some and may be inconvenient for people with work or caring commitments.^6^ Uptake of face-to-face groups is lower among younger people.^7-9^ Digital delivery may be a more attractive alternative for some. Digital services can be effective in achieving weight loss and dietary changes in the wider population ^10, 11^ and among people with T2D.^12, 13^ Digital services have been tested head-to-head against face-to-face services for other health conditions and shown to be as effective.^10, 14^ However, to our knowledge, although digital interventions for T2D prevention were associated with positive changes in weight of similar magnitude to face-to-face delivery,^15-18^ there has been no direct comparison of face-to-face and digital DPPs.

Lower socioeconomic status and black or Asian ethnicity were associated with poorer outcomes in the face-to-face English DPP.^8, 19^ A systematic review of digital services based on the US DPP model reported that, compared to the original face-to-face model, the digital cohorts had a higher BMI, less ethnic diversity, higher educational achievement and were predominantly female,^17^ raising concerns that digital delivery has the potential to worsen existing inequalities in DPP outcomes.

The National Health Service Diabetes Prevention Programme (NHS DPP), “Healthier You” was offered to people in England at high risk of developing T2D, starting in 2016.^20^ Initially, the service was only delivered through group-based face-to-face sessions, because of the limited evidence for a digital alternative. A digital mode of delivery was developed by NHS England as a pilot in 2017-18, to support a decision on whether to proceed with national roll-out of a digital alternative.^21, 22^ The underlying content and approaches of the digital and face-to-face programmes were similar: both services were commissioned by NHS England from external providers, using service specifications which set out comparable requirements in terms of behaviour change and self-management content.

The contemporaneous delivery of these two modes of delivery of the same programme provided a unique opportunity to compare the effectiveness of face-to-face and digital delivery. Due to the stronger evidence base for face-to-face delivery, we were interested in determining if outcomes in digital delivery were at least as good. We were also interested to observe the impact of the digital service on inequalities. The aims were: (1) evaluate whether change in weight from baseline to 6 months (primary) and 12 months in digital delivery was no worse than that in face-to-face delivery, and (2) evaluate whether patient characteristics were associated with differential changes in weight between the digital and face-to-face delivery.

The protocol was pre-registered on the Open Science Framework 14 July 2021.^23^

## RESEARCH DESIGN AND METHODS

This study was a retrospective observational cohort study, using patient-level data collected by NHS DPP service providers.

### Study interventions and populations

The NHS DPP was developed to encourage healthy eating, weight loss and increased exercise in adults at high risk of developing T2D, defined as having nondiabetic hyperglycaemia (NDH) (HbA1c 42–47mmol/mol [6.0–6.4%] or fasting plasma glucose (FPG, 5.5-6.9mmol/L).^8, 24, 25^ The NHS DPP programme was developed by NHS England, supported by an Expert Reference Group,^26^ and incorporated strategies shown to be effective at influencing behaviour.^27-29^ The programme was gradually rolled out across England from 2016, with national coverage by 2018, accessed by referral from general practice. Until 2020, the primary delivery of the NHS DPP was through group-based face-to-face sessions. Once referred, participants were invited to attend an initial assessment, followed by at least thirteen group-based face-to-face sessions over 9-12 months, delivered by one of four (later five) service providers. Content of the programme varied across the providers but, in general, comprised behaviour change around diet, weight loss and increased exercise with regular group education and exercise sessions.^28, 30^

A digital pilot was offered in nine areas in England, by one of five service providers. The referral procedure was the same as that for face-to-face delivery. The digital pilot was delivered via two delivery models: (i) digital-only - in areas where face-to-face delivery had not yet been rolled out, participants were only offered digital, (ii) digital-choice - four areas offered patients a choice between digital and face-to-face delivery. Participants from the digital-only and choice areas were separately compared with participants in the face-to-face cohort and are subsequently referred to as the digital-only and digital-choice cohorts. Our main interest was in the comparison of digital-only with face-to-face delivery due to the built-in selection in the digital-choice group.

To maximise comparability, we included data from face-to-face delivery contemporaneous with the period of digital recruitment (1^st^ December 2017 and 31^st^ December 2018).

The inclusion/exclusion criteria for both programmes are shown in Table 1.

**Table 1.** Inclusion/exclusion criteria for the face-to-face and digital pilot programmes

|  |  |
| --- | --- |
| Inclusion criteria | 1) Aged 18 or over<br>2) Registered with a GP participating in the programme<br>3) NDH defined as HbA1c of 42–47 mmol/mol (6.0–6.4%) or fasting plasma glucose level (FPG) of 5.5–6.9 mmol/l, measured up to 12 months prior to referral. |
| Exclusion criteria | 1) Pregnancy<br>2) Diagnosis of diabetes<br>3) Digital programme only: underweight, defined as BMI<18.5.<br>To equalise the exclusion criteria between the two groups, we excluded individuals in the face-to-face group who were underweight at the first session. |

### Patient and site characteristics

Available data were age at referral, sex, ethnicity, socioeconomic deprivation (defined by the English indices of deprivation 2015 associated with the lower layer super output area derived from the individual’s postcode, grouped into quintiles), weight in kg, HbA1c in mmol/mol and BMI in kg/m^2^. The site in which the participant resided was described via health-administration geographical areas: Clinical Commissioning Group (CCG), local General Practice led statutory bodies which commission health services; and Sustainability and Transformation Partnership (STP), super-imposed larger geographical footprints responsible for planning for the long term needs of populations. Each STP commissioned a single provider and CCGs managed the local implementation of referrals from General Practice.

### Outcome measures

The pre-specified co-primary outcomes were (i) change in weight and (ii) change in HbA1c, at 6 (primary) and 12 months.

#### Weight

Weight was objectively recorded using pre-calibrated equipment at group sessions (face-to-face), in General Practice, or at home using equipment supplied by the provider which automatically uploaded the recorded weight. There were no self-reported weight measures. The same mode of measurement was used for baseline and follow-up observations. We defined baseline weight as that measured at the first intervention session attended (face-to-face) or registration (digital), 6-month weight as that closest to 6 months after baseline (within 4-8 months), and 12-month weight as that closest to 12 months after baseline (within 8-14 months).

The pre-specified non-inferiority margins for change in weight were determined by the Expert Reference Group^26^ as 1kg at 6 months and 0.7kg at 12 months. For example, if change in weight at 6 months in the face-to-face delivery was no greater than 1kg more than in digital delivery, digital was deemed non-inferior.

Weight in face-to-face delivery was measured from individuals who were participating in the programme at the time. Hence, estimated changes in weight apply only to those who were still enrolled. In the digital pilot, all individuals who registered were invited to provide 6- and 12-month data, regardless of whether they were still enrolled.

#### HbA1c

Use of HbA1c measures in this study were problematic. Firstly, HbA1c measurement differed across the two delivery modes. In face-to-face, point-of-care tests were used. In digital, four providers used venous blood tests and one used a home test. Previous work has suggested point-of-care tests tend to give lower values than venous methods.^31^ We were uncertain how much impact differences in measurement would have. Secondly, only 21.3% of the face-to-face cohort provided baseline and 6-month measures, making the matching pool too small to identify the required number of matches. Thirdly, a non-inferiority limit margin could not be pre-specified for change in HbA1c, due to lack of international guidance on a clinically important change among people in the NDH range. Given these data limitations, we have deviated from the protocol and omitted HbA1c findings.

### Statistical analyses

#### Matching

Individuals from the digital pilot were matched to individuals from face-to-face delivery to account for confounding, using a similar approach to previous studies.^32-34^ We matched on sex, age (withing 3 years), ethnicity (categorised as White/Mixed/Black/Asian/Other) and deprivation quintile. If one or more of these were missing, the participant was excluded from the main analysis. Within these constraints, matches from the face-to-face cohort were randomly chosen without replacement. We sought up to five matches from face-to-face per digital observation. As there were differences in baseline characteristics between the digital-only and digital-choice cohorts, matching was performed separately in these groups, and at different time points.

#### Primary analysis – matched data

In the matched cohorts, mixed effects linear regression modelling was used to compare change in weight from baseline to 6 months between the face-to-face cohort and each digital cohort. A binary indicator variable (face-to-face/digital) was included to convey the estimated adjusted difference in mean change in weight between delivery types, with face-to-face as the reference group. The 95% confidence interval for these adjusted differences was obtained and the bounds determined if non-inferiority had been demonstrated. The model adjusted for the matching variables and timing of the outcome measure (months from baseline) as fixed effects, and CCG nested within STP as random effects to account for variation across sites. Changes at 12 months were analysed in the same way.

#### Supplementary analyses

We re-ran the models using (i) regression adjustment to account for confounding, instead of matching (using the full cohort, with a larger sample size), (ii) multiple imputation of missing values (Supplemental File) and (iii) making a range of plausible assumptions about weight change in individuals with missing values.^35^

The whole (un-matched) cohort was used to investigate inequalities in weight change, because a matched dataset is unsuitable for estimating associations with covariates using in the matching process.^36^ The same mixed effects linear regression modelling approach was taken, with an additional interaction term between the cohort indicator variable (face-to-face/digital) and each of age, sex, ethnicity, deprivation and baseline weight. Each interaction was analysed in a distinct model.

#### Sensitivity analysis

Sensitivity analyses were undertaken exploring the effect of different outcome measurement time points. (Supplemental File).

Analyses were performed in Stata version 14.^37^

## Sample size

Although the sample size was predetermined by the available data, we performed sample size calculations and determined that the cohort sizes were sufficient to detect the pre-specified non-inferiority limit at 6 months. More information is given in the protocol ^23^ and the Supplemental File.

## RESULTS

### Baseline characteristics

There were 65051 individuals who attended a first session in the face-to-face cohort, 1776 individuals who registered in the digital-only cohort and 1412 in the digital-choice cohort. Compared to face-to-face, the digital-only cohort was, on average, younger, from more deprived communities, with a higher proportion of people from an Asian ethnicity, higher baseline weight, BMI and HbA1c. The distribution of sex was similar. Compared to face-to-face, the digital-choice cohort was on average, younger, had a higher proportion of males, from more deprived communities, higher baseline weight. The distribution of ethnicity and BMI were similar. Those in the digital-choice and digital-only cohorts had a similar mean age, baseline weight and baseline HbA1c, but there was a higher proportion of males, individuals of White ethnicity and individuals in the most deprived quintile in digital-choice. (Table 2)

**Table 2.**
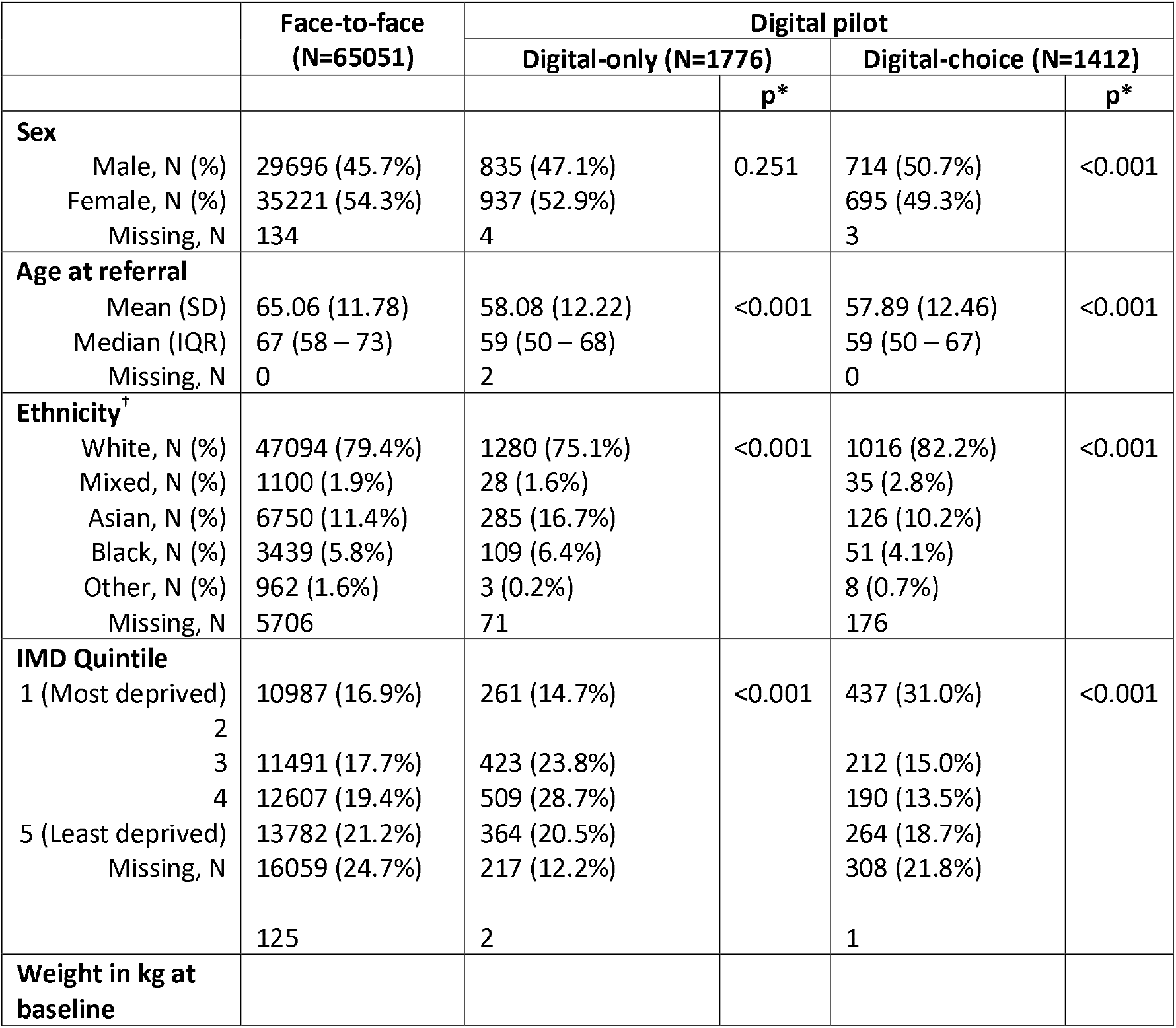
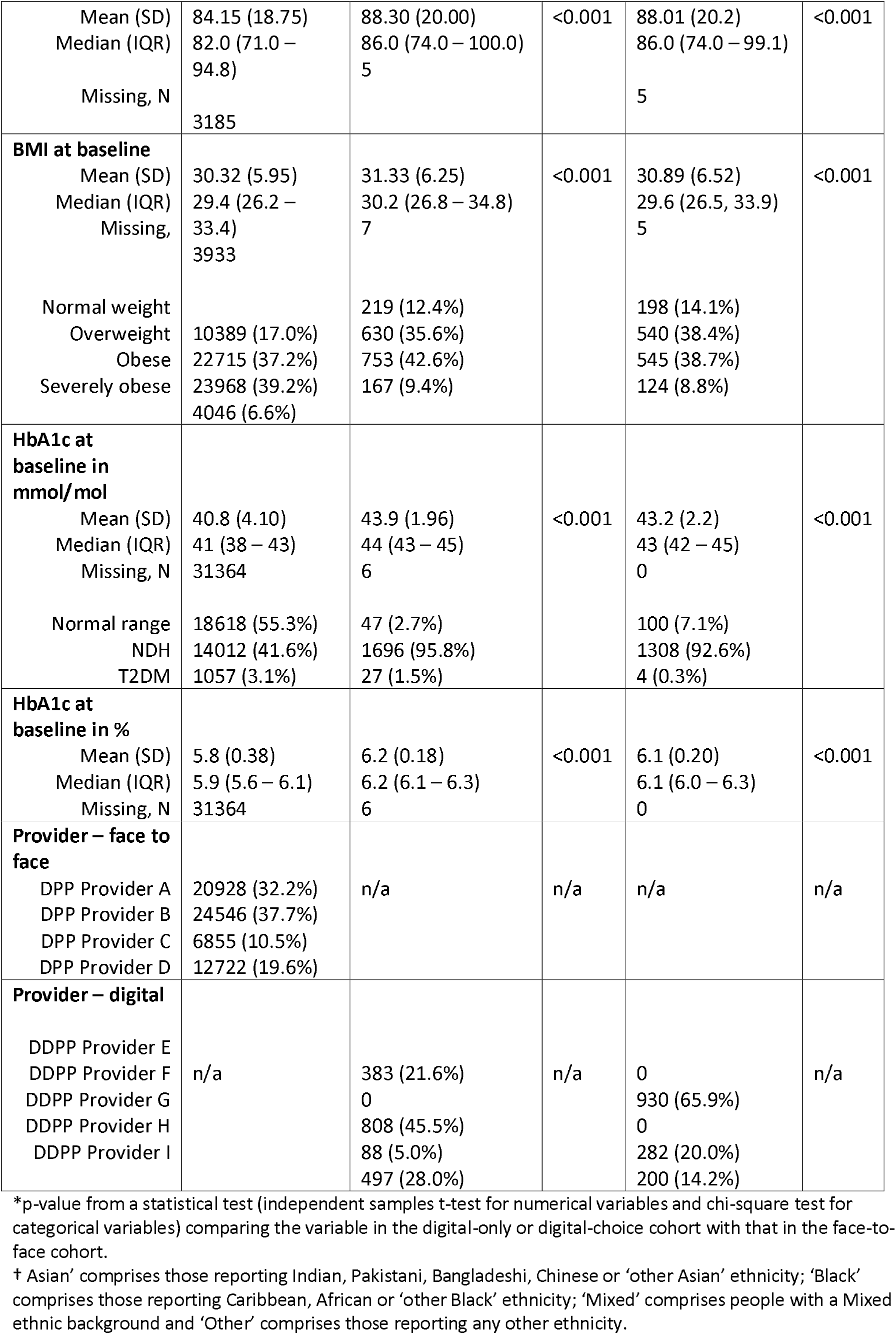
Baseline characteristics of participants in the face-to-face cohort, digital-only cohort and digital-choice cohort

### Outcome summary

The mean changes in weight at 6 months were: −3.05kg (95% CI: −3.38, −2.73) digital-only, −3.79kg (− 4.16, −3.43) digital-choice and −2.85kg (−2.89, −2.81) face-to-face, where a negative value indicates a reduction from baseline weight. Hence, all modes of delivery saw a mean reduction in weight at 6 months. Changes at 12 months were also negative, indicating reductions in weight. (Table 3).

**Table 3.**
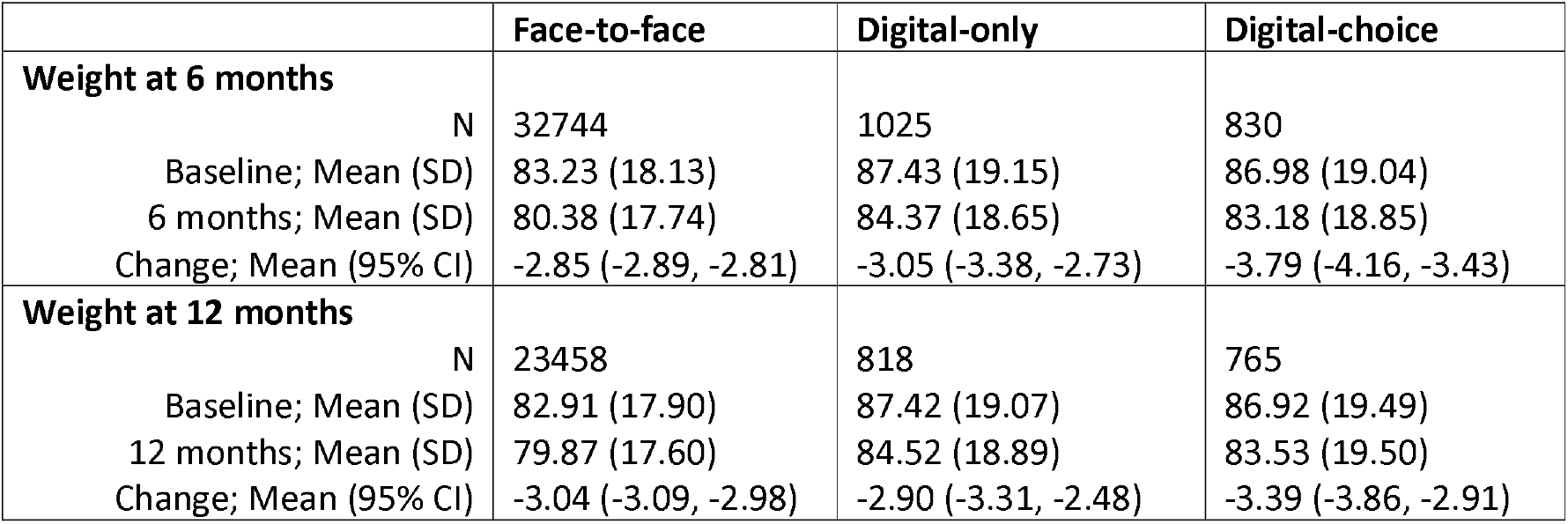
Summary of weight outcome measures of participants in the face-to-face cohort and digital cohorts

|  | Face-to-face | Digital-only | Digital-choice |
| --- | --- | --- | --- |
| **Weight at 6 months** |  |  |  |
| N | 32744 | 1025 | 830 |
| Baseline; Mean (SD) | 83.23 (18.13) | 87.43 (19.15) | 86.98 (19.04) |
| 6 months; Mean (SD) | 80.38 (17.74) | 84.37 (18.65) | 83.18 (18.85) |
| Change; Mean (95% CI) | -2.85 (-2.89, -2.81) | -3.05 (-3.38, -2.73) | -3.79 (-4.16, -3.43) |
| **Weight at 12 months** |  |  |  |
| N | 23458 | 818 | 765 |
| Baseline; Mean (SD) | 82.91 (17.90) | 87.42 (19.07) | 86.92 (19.49) |
| 12 months; Mean (SD) | 79.87 (17.60) | 84.52 (18.89) | 83.53 (19.50) |
| Change; Mean (95% CI) | -3.04 (-3.09, -2.98) | -2.90 (-3.31, -2.48) | -3.39 (-3.86, -2.91) |

Largely due to drop-out from the programme, baseline and 6-month weight data were only available for 50.3% (face-to-face), 57.7% (digital-only) and 58.8% (digital-choice) of participants. Across face-to-face and digital-only delivery, data was more likely to be missing from participants who were younger, from ethnic minorities, with higher baseline weights. Higher deprivation was associated with being missing in face-to-face and digital-choice delivery.^35^

Matching rates were high across all matched datasets, ranging from 98.1% to 99.4%. The matched samples were very similar in terms of the matched variables (Tables S1-S2). The mean weight was approximately 2kg higher in the digital-only cohort than face-to-face, but the mean BMI was similar.

Table 4 shows the results of the mixed effects linear regression analysis comparing change in weight at 6 months and 12 months between the digital and face-to-face cohorts.

**Table 4.** Regression analyses comparing change in weight from baseline to 6 months and 12 months between the face-to-face cohort and the digital cohorts

|  |  | N | B* | 95% CI | p-value |
| --- | --- | --- | --- | --- | --- |
| **Digital only vs Face-to Face** | **Weight at 6 months<sup>†</sup>** | 5726 | -0.284 | (-0.712, 0.144) | 0.194 |
|  | **Weight at 12 months<sup>†</sup>** | 4589 | -0.466 | (-1.068, 0.137) | 0.130 |
| **Digital choice vs Face-to-Face** | **Weight at 6 months<sup>†</sup>** | 4504 | -1.165 | (-1.841, -0.489) | 0.001 |
|  | **Weight at 12 months<sup>†</sup>** | 4040 | -1.009 | (-1.738, -0.280) | 0.007 |

\*Coefficient quantifies the difference in mean change between the face-to-face and digital cohort, using the face-to-face cohort as the reference group
† Model adjusts for age at referral, sex, ethnicity (white/mixed/black/Asian/other), IMD quintile, time since baseline (in months) as fixed effects and CCG (site) nested within STP as random effects

### Comparison of digital-only and face-to-face

#### Change in weight at 6 months

962 digital-only participants were matched to 4764 face-to-face participants. At 6 months individuals in the digital-only cohort lost, on average, 0.284kg more weight than the face-to-face cohort. Non-inferiority of digital-only delivery was demonstrated as the upper limit of the 95% confidence interval (−0.712, 0.144kg) was less than the non-inferiority limit of 1.0kg.

#### Change in weight at 12 months

770 digital-only participants were matched to 3819 face-to-face participants. At 12 months individuals in the digital-only cohort lost, on average, 0.466kg more weight than the face-to-face cohort. Non-inferiority of digital-only delivery was demonstrated as the upper limit of the 95% confidence interval (−1.068, 0.137) was less than the non-inferiority limit of 0.7kg.).

#### Interaction effects

Whilst both sexes, on average, lost weight on both programmes, males lost more weight on the face-to-face programme compared to the digital-only programme (although this was not statistically significant) and females lost more weight on the digital-only programme (Table 5). There was strong evidence of a difference in weight loss between males and females across the two programmes (p<0.001).

**Table 5.**
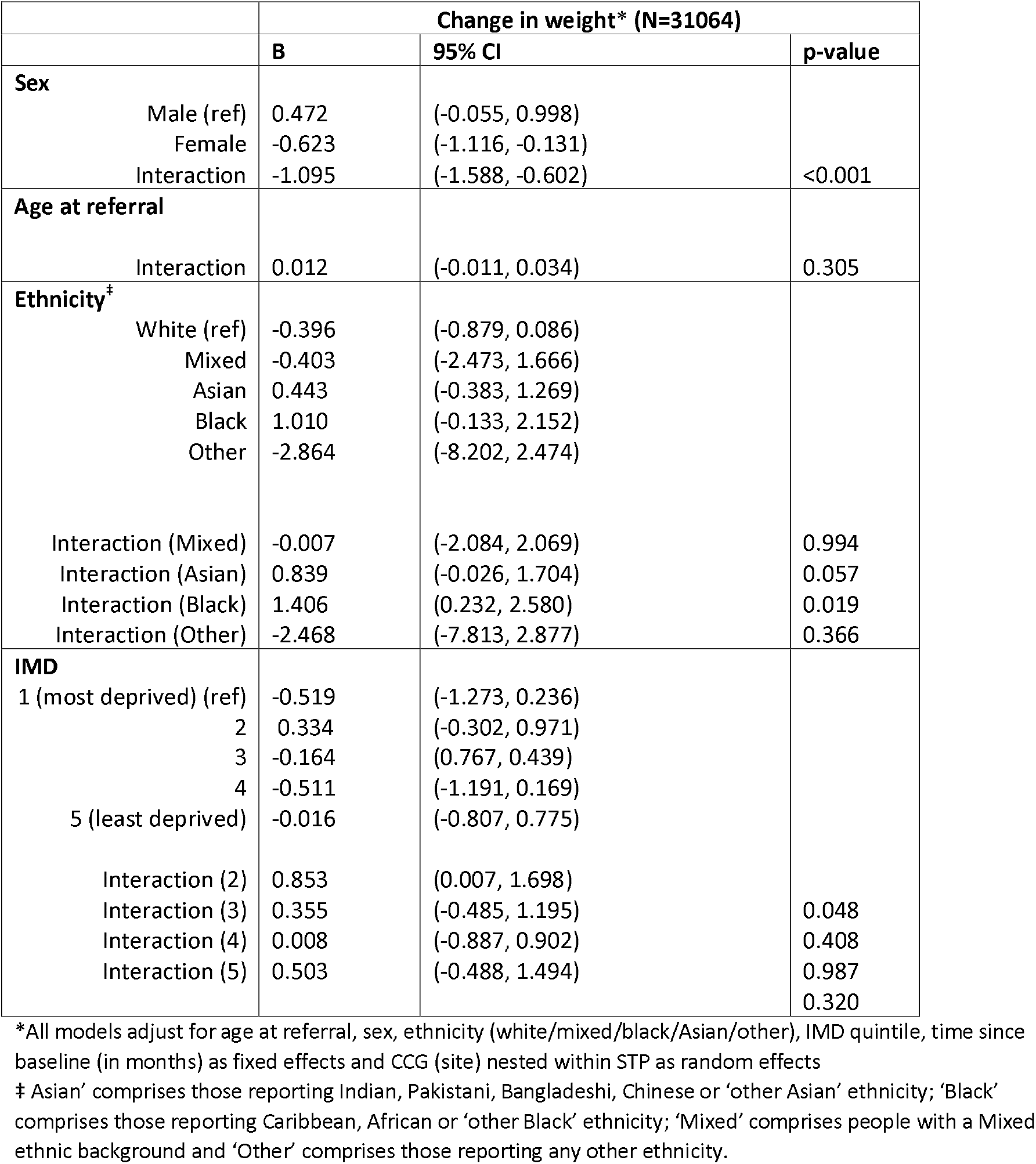
Interaction analyses assessing differential change in weight from baseline to 6 months between the face-to-face cohort and the digital-only cohort. Subgroup effects are shown, where relevant

How to read the table – using example of sex: Males, in the face-to-face cohort lost, on average, 0.472kg more than males in the digital-only cohort, and females, in the face-to-face cohort lost, on average, 0.623kg less than females in the digital-only cohort. The interaction effect of −1.095 is the difference between the effect in females and the effect in males. As the interaction effect is statistically significant (p<0.001), there is strong evidence that the difference in change in weight between the digital-only and face-to-face cohorts varies by sex. Furthermore, the 95% confidence intervals around the subgroup effects provide evidence that females lose more weight on the digital programme but, as the 95% confidence interval around the effect for males contains 0, there is insufficient evidence to suggest males lost more weight on one programme than the other.

Evidence of a difference across ethnic groups was seen. All ethnic groups, on average, lost weight on both programmes and White individuals lost more weight than other ethnic groups on both programmes (data not shown). Although the subgroup effects were not statistically significant, the point estimates suggested White individuals lost more weight on the digital programme than the face-to-face programme whilst Asian and Black individuals lost more weight on the face-to-face programme than the digital programme. However, the interaction effect between White and Black ethnic groups was 1.406 (95% CI: 0.232, 2.580), suggesting the difference in weight loss between White and Black individuals was statistically significantly larger in the digital programme than the face-to-face programme. The interaction effect between White and Asian individuals was 0.839 (95% CI: −0.026, 1.704), suggesting that the difference in weight loss between White and Asian individuals may be larger in the digital programme than the face-to-face programme, but this was not statistically significant. Results should be viewed with caution however due to the small number of Black and Asian individuals in the digital cohort. No interpretation of the results regarding the Mixed and Other ethnic groups is attempted due to the small sample size of these groups.

There was no evidence of an interaction between age and delivery mode, nor between the least deprived quintile and most deprived quintiles.

### Comparison of digital-choice and face-to-face

Table 4 shows the results of the mixed effects linear regression analysis comparing change in weight at 6 and 12 months between the digital-choice and face-to-face cohorts.

#### Change in weight at 6 months

758 digital-choice participants were matched to 3746 face-to-face participants. At 6 months, individuals in the digital-choice cohort lost, on average, 1.165kg more weight than the face-to-face cohort. Non-inferiority was demonstrated as the upper limit of the 95% confidence interval (−1.841, − 0.489kg) was less than 1.0kg.

#### Changes in weight at 12 months

687 digital-choice participants were matched to 3353 face-to-face participants. At 12 months, individuals in the digital-choice cohort lost, on average, 1.009kg more weight than the face-to-face cohort. Non-inferiority was demonstrated as the upper limit of the 95% confidence interval (−1.738, − 0.280) was less than 0.7kg (Table 4).

#### Interaction effects

Interactions between delivery mode and age, ethnicity and deprivation quintile on change in weight at 6 months were observed. More information is given in the Supplemental File (Table S3).

### Additional analyses

Full results from all additional analyses are shown in the Supplemental File (Tables S4-S12). Results were similar to the main analysis and conclusions were the same.

## DISCUSSION

Participants lost weight, on average, whether the DPP was delivered face-to-face or digitally and whether or not they had a choice. After accounting for differences in sex, age, deprivation and ethnicity, weight loss after digital delivery (in both those who were and were not offered a choice) was non-inferior to that in face-to-face delivery at both 6 and 12 months. Furthermore, whilst differences in weight loss were similar for face-to-face and digital-only delivery, weight loss was greater when participants were offered a choice and chose digital, particularly at 6 months. These results were robust to several analysis approaches including matching, regression adjustment and multiple imputation.

Previous research has suggested digital delivery can be effective,^10, 15-18^ but this study is novel in offering a head-to-head comparison. The unadjusted weight loss was similar to that reported elsewhere: mean weight loss at 12 months of 3.3-3.6kg in the face-to-face service,^8, 19^, and 3.1kg (95% CI: 2.8, 3.4) in the digital service.^38^ We have gone one step further, showing that, when compared directly, taking account of differences in site and personal characteristics, digital delivery was non-inferior to face-to-face, among participants.

Non-diabetic hyperglycaemia is more prevalent among older people and those living in deprived areas,^39^ and there is a risk of health interventions exacerbating such health inequalities.^40^. Participation in the English DPP is lower among people from deprived areas, younger people and those with disabilities, but participation of ethnic minorities is good.^41^. Therefore, an important consideration is whether a DPP worsens existing inequalities. In this study we found that, across sex, age, ethnicity and deprivation, all population categories lost weight, on average, regardless of delivery mode. Whether the service was delivered face-to-face or digitally, white lost more weight than black individuals, and the difference in weight loss between white and black was greater in digital delivery compared to face-to-face. Women lost more weight via digital delivery than face-to-face, and men lost more on face-to-face. People from the most deprived areas had similar weight loss on both programmes. Overall, this suggests that targeting of disadvantaged groups would be beneficial for the DPP, but digital delivery is unlikely to widen disparities, particularly if people are offered a choice.

Strengths of the study include that it offers a rare opportunity to compare a DPP that was delivered face-to-face and digitally contemporaneously to similar populations, offering similar content and following service specification from NHS England which set out comparable requirements in terms of behaviour change and self-management. Although a randomised controlled trial would have been the optimal way to compare delivery modes, this study provides a quicker and cheaper approach. This study had a large sample size and access to data from all participants who took part during the time-period of interest.

The limitations arise from differences in the organisation of the programmes and how data was collected. Firstly, different ways of measuring HbA1c were used which precluded direct comparison of HbA1c change between delivery modes. However, HbA1c changes tend to be strongly associated with weight change.^8^ Secondly, outcome data were collected only from participating individuals in the face-to-face delivery, whilst all individuals on the digital pilot were invited to provide outcome data. Any bias arising from this difference is likely to favour face-to-face delivery, assuming that drop-out is related to lack of success in losing weight. As we found non-inferiority of digital delivery this is not problematic. Our analysis, like most DPP studies,^4, 8, 19, 38^ had data missing not at random, from people who did not complete the course, so these results apply to completers rather than all who were referred. Elsewhere we have mitigated this by reporting a supplementary analysis imputing plausible outcomes.^35^ Baseline weight was higher in the digital only cohort than the face-to-face cohort, even after matching, and we have minimised the effect of this by modelling change in weight. Lastly, we were only able to account for differences in sex, age, ethnicity, deprivation and site: the results may be biased by unmeasured confounding.

Future research is warranted comparing effectiveness of digital and face-to-face delivery of DPPs in the total referred population, rather than, as our data allowed, in participants. Research is also needed to compare drop-out rates and achieved dose levels between digital and face-to-face delivery, both across the population and in sub-groups to further understanding of the potential for reducing health inequity.

## CONCLUSION

Among patients at risk of type 2 diabetes who complete a diabetes prevention programme, digital delivery can achieve weight loss at least as high as face-to-face group delivery, demonstrating that the same content can be delivered in alternative ways without loss of impact. Where patients are offered a choice of digital or face-to-face, those that choose digital have better weight loss outcomes than those on face-to-face who were offered no choice. In response to this and other evidence, since 2022, patients starting the NHS DPP have been offered a choice of face-to-face group-based delivery or digital delivery.^20^ Whilst all patient groups, on average, lost weight on both programmes, offering digital delivery may benefit some groups more than others.

## Supporting information

STROBE checklist

## Data Availability

The data that support the findings of this study were used under license from NHS England for the current study only and are not publicly available.

## ACKNOWLEDGEMENTS

We are grateful to members of the Diabetes Prevention Programme Expert Reference Group^26^ who provided valuable feedback on the design and execution of this research. We would like to thank Professor Evan Kontopantelis for advice about methodology for matching studies and Professor Peter Bower who provided valuable feedback on the research design and during the manuscript preparation.

## ETHICS COMMITTEE APPROVAL

The study was pre-registered on the Open Science Framework repository (DOI:<u>10.17605/OSF.IO/A9PBW</u>). The research was approved by the North-West Greater Manchester East NHS Research Ethics Committee (Reference: 17/NW/0426, 01/08/17, amended 29/09/20).

## AUTHORS’ CONTRIBUTIONS

AM designed the study, performed the statistical analyses, interpreted the results and prepared the manuscript. MH designed the study, supervised the research conduct and contributed to interpretation of results. EB contributed to the study design and interpretation of results. BM contributed to the interpretation of results. EM contributed to the study design and interpretation of results. JV contributed to the study design and interpretation of results. SC designed the study, secured funding, supervised the research conduct and contributed to interpretation of results. All authors read and approved the final manuscript.

