## Supplemental file for "The effectiveness of digital delivery versus group-based face-to-face delivery of the English National Health Service Diabetes Prevention Programme: a non-inferiority retrospective cohort comparison study"

**The comparative effectiveness of a diabetes prevention programme delivered face-to-face and by digital methods: a non-inferiority retrospective cohort comparison study**

**Online-Only Supplemental Material**

**Sample size justification**

The original sample size calculation is shown in the online pre-registration on the Open Science Framework repository (DOI:[10.17605/OSF.IO/A9PBW](https://doi.org/10.17605/OSF.IO/A9PBW)). In the protocol, we planned to match each digital observation to up to 5 face-to-face observations for both the change in weight and HbA1c analyses. However, we subsequently decided to use up to only three matches per digital observation for the HbA1c analyses due high rates of missing baseline HbA1c in the face-to-face cohort. We also used the 12-month non-inferiority margin instead of the 6-month margin for the change in weight calculations. The revised sample size calculations are presented here.

In the pilot digital data, there are 1025 individuals from a digital-only area who have both a baseline and 6-month weight measure. The pooled standard deviation of change in weight from baseline to 6 months for eligible participants in the digital-only and face-to-face cohorts is 3.89kg. Assuming a 5/1 ratio of F2f participants to digital participants, to detect a non-inferiority margin of 1kg in a two-sample t-test with 90% power, a significance level of 0.05 and a population standard deviation of 3.89, 156 participants in the digital-only group are required.

Also in the pilot digital data, there are 955 individuals from a digital-only area who have both a baseline HbA1c and 6 month HbA1c measure. The pooled standard deviation of change in HbA1c from baseline to 6 months for eligible participants in the digital and face-to-face cohorts is 4.43mm/mol. Whilst we have not specified an non-inferiority margin for change HbA1c, assuming a 3/1 ratio of F2f participants to digital participants, with 955 in the digital-only group and 2865 in the F2f group, 90% power, a significance level of 0.05 and a population standard deviation of 4.43, a two-sample t-test can detect a non-inferiority limit of 0.49mm/mol. The original sample size calculation determined the test could detect a non-inferiority limit of 0.49mm/mol with a 5/1 ratio and all other factors kept the same.

**Summary of baseline characteristics in the matched cohorts**

Table S1 Baseline characteristics of participants in matching cohorts for the change in weight at 6m analyses comparing digital-only and digital-choice with face-to-face

|  | **Digital only vs face-to-face**  **Matching rate: 98.8%** | | **Digital choice vs face-to-face Matching rate: 99.2%** | |
| --- | --- | --- | --- | --- |
|  | **Digital only (N=962)** | **Face-to-face (N=4764)** | **Digital choice (N=758)** | **Face-to-face (N=3746)** |
| **Sex**  Male, N (%) Female, N (%) | 439 (45.6%)  523 (54.4%) | 2176 (45.7%)  2588 (54.3%) | 371 (48.9%)  387 (51.1%) | 1840 (49.1%)  1906 (50.9%) |
| **Age at referral**  Mean (SD) Median (IQR) | 60.0 (11.7)  61 (52, 69) | 60.8 (11.5)  62 (53, 70) | 60.0 (11.3)  61 (53, 69) | 61.0 (11.1)  62 (54, 69) |
| **Ethnicity***  White, N (%)  Mixed, N (%)  Asian, N (%)  Black, N (%)  Other, N (%) | 755 (78.5%)  12 (1.25%)  138 (14.4%)  55 (5.72%)  2 (0.21%) | 3760 (78.9%)  48 (1.01%)  678 (14.2%)  270 (5.67%)  8 (0.17%) | 640 (84.4%)  16 (2.11%)  75 (9.89%)  23 (3.03%)  4 (0.53%) | 3188 (85.1%)  62 (1.66%)  365 (9.74%)  115 (3.07%)  16 (0.43%) |
| **IMD Quintile**  1 (Most deprived)  2  3  4  5 (Least deprived) | 142 (14.8%)  214 (22.3%)  288 (29.9%)  196 (20.4%)  122 (12.7%) | 695 (14.6%)  1066 (22.4%)  1421 (29.8%)  978 (20.5%)  604 (12.7%) | 178 (23.5%)  101 (13.3%)  96 (12.7%)  175 (23.1%)  208 (27.4%) | 870 (23.2%)  498 (13.3%)  480 (12.8%)  866 (23.1%)  1032 (27.6%) |
| **Weight in kg at baseline**  Mean (SD) Median (IQR) | 87.6 (19.2)  85.0 (73.9, 99.0) | 85.9 (19.4)  83.6 (72.1, 96.4) | 86.9 (19.1)  84.8 (73.9, 98.0) | 87.2 (19.4)  85.4 (73.4, 98.2) |
| **BMI at baseline**  Mean (SD) Median (IQR) | 31.1 (6.00)  30.1 (26.6, 34.5) | 30.7 (6.1)  29.8 (26.5, 33.8) | 30.5 (6.1)  29.3 (26.2, 33.4) | 30.9 (6.2)  30.0 (26.6, 34.1) |

* Asian’ comprises those reporting Indian, Pakistani, Bangladeshi, Chinese or ‘other Asian’ ethnicity; ‘Black’ comprises those reporting Caribbean, African or ‘other Black’ ethnicity; ‘Mixed’ comprises people with a Mixed ethnic background and ‘Other’ comprises those reporting any other ethnicity.

Table S2 Baseline characteristics of participants in matching cohorts for the change in HbA1c at 6m analyses comparing digital-only and digital-choice with face-to-face

|  | **Digital only vs face-to-face**  **Matching rate: 95.1%** | | **Digital choice vs face-to-face**  **Matching rate: 96.9%** | |
| --- | --- | --- | --- | --- |
|  | **Digital only (N=863)** | **Face-to-face (N=2529)** | **Digital choice (N=738)** | **Face-to-face (N=2179)** |
| **Sex**  Male, N (%) Female, N (%) | 402 (46.6%)  461 (53.4%) | 1173 (46.4%)  1356 (53.6%) | 361 (48.9%)  377 (51.1%) | 1071 (49.2%)  1108 (50.9%) |
| **Age at referral**  Mean (SD) Median (IQR) | 59.8 (11.1)  61 (52, 68) | 61.0 (11.0)  62 (54, 69) | 60.3 (11.1)  61 (53, 69) | 61.4 (10.9)  63 (54, 70) |
| **Ethnicity***  White, N (%)  Mixed, N (%)  Asian, N (%)  Black, N (%)  Other, N (%) | 662 (77.1%)  7 (0.81%)  127 (14.7%)  63 (7.30%)  1 (0.12%) | 1984 (78.5%)  17 (0.67%)  346 (13.7%)  179 (7.08%)  3 (0.12%) | 629 (85.2%)  12 (1.63%)  68 (9.21%)  26 (3.52%)  3 (0.41%) | 1873 (86.0%)  28 (01.28%)  196 (8.99%)  76 (3.49%)  6 (0.28%) |
| **IMD Quintile**  1 (Most deprived)  2  3  4  5 (Least deprived) | 135 (15.6%)  192 (22.1%)  251 (29.1%)  175 (20.3%)  111 (12.9%) | 391 (15.5%)  557 (22.0%)  735 (29.1%)  519 (20.5%)  327 (12.9%) | 182 (24.7%)  106 (14.4%)  91 (12.3%)  165 (22.4%)  194 (26.3%) | 531 (24.4%)  312 (14.3%)  269 (12.4%)  490 (22.5%)  577 (26.5%) |
| **Weight in kg at baseline**  Mean (SD) Median (IQR) | 87.9 (19.7)  85.0 (73.8, 99.2) | 85.8 (19.5)  83.2 (71.8, 97.3) | 86.4 (19.2)  84.0 (72.7, 97.5) | 86.5 (19.5)  84.2 (72.2, 98.0) |
| **BMI at baseline**  Mean (SD) Median (IQR) | 31.3 (6.17)  30.1 (26.7, 34.8) | 30.7 (6.2)  29.7 (26.5, 33.8) | 30.3 (5.97)  29.2 (26.0, 33.3) | 30.8 (6.1)  29.8 (26.3, 34.2) |

* Asian’ comprises those reporting Indian, Pakistani, Bangladeshi, Chinese or ‘other Asian’ ethnicity; ‘Black’ comprises those reporting Caribbean, African or ‘other Black’ ethnicity; ‘Mixed’ comprises people with a Mixed ethnic background and ‘Other’ comprises those reporting any other ethnicity.

Table S3 Baseline characteristics of participants in matching cohorts for the change in weight at 12m analyses comparing digital-only and digital-choice with face-to-face

|  | **Digital only vs face-to-face**  **Matching rate: 99.4%** | | **Digital choice vs face-to-face**  **Matching rate: 98.1%** | |
| --- | --- | --- | --- | --- |
|  | **Digital only (N=770)** | **Face-to-face (N=3819)** | **Digital only (N=687)** | **Face-to-face (N=3353)** |
| **Sex**  Male, N (%) Female, N (%) | 366 (47.5%)  404 (52.5%) | 1809 (47.4%)  2010 (52.6%) | 334 (48.6%)  353 (51.4%) | 1633 (48.7%)  1720 (51.3%) |
| **Age at referral**  Mean (SD) Median (IQR) | 60.9 (11.0)  62 (53, 70) | 61.7 (11.0)  63 (54, 70) | 60.1 (11.1)  61 (53, 68) | 61.3 (10.9)  62 (54, 69) |
| **Ethnicity***  White, N (%)  Mixed, N (%)  Asian, N (%)  Black, N (%)  Other, N (%) | 617 (80.1%)  6 (0.78%)  103 (13.4%)  43 (5.58%)  1 (0.13%) | 3070 (80.4%)  28 (0.73%)  506 (13.3%)  213 (5.58%)  2 (0.05%) | 583 (84.9%)  17 (2.47%)  62 (9.02%)  22 (3.20%)  3 (0.44%) | 2886 (86.1%)  54 (1.61%)  294 (8.77%)  106 (3.16%)  13 (0.39%) |
| **IMD Quintile**  1 (Most deprived)  2  3  4  5 (Least deprived) | 95 (12.3%)  169 (22.0%)  242 (31.4%)  158 (20.5%)  106 (13.8%) | 468 (12.3%)  841 (22.0%)  1199 (31.4%)  785 (20.6%)  526 (13.8%) | 161 (23.4%)  88 (12.8%)  89 (13.0%)  157 (22.9%)  192 (28.0%) | 780 (23.3%)  422 (12.6%)  441 (13.2%)  722 (23.0%)  938 (28.0%) |
| **Weight in kg at baseline**  Mean (SD) Median (IQR) | 87.6 (19.1)  85.4 (73.5, 100.0) | 85.0 (19.2)  82.8 (71.5, 95.8) | 86.8 (19.6)  85.0 (73.0, 98.0) | 86.4 (19.7)  83.6 (72.8, 97.4) |
| **BMI at baseline**  Mean (SD) Median (IQR) | 31.1 (5.94)  30.1 (26.7, 34.5) | 30.3 (6.1)  29.8 (26.5, 33.8) | 30.3 (6.0)  29.3 (26.1, 33.3) | 30.6 (6.1)  29.6 (26.5, 33.7) |

* Asian’ comprises those reporting Indian, Pakistani, Bangladeshi, Chinese or ‘other Asian’ ethnicity; ‘Black’ comprises those reporting Caribbean, African or ‘other Black’ ethnicity; ‘Mixed’ comprises people with a Mixed ethnic background and ‘Other’ comprises those reporting any other ethnicity.

Table S4 Baseline characteristics of participants in matching cohorts for the change in HbA1c at 12m analyses comparing digital-only and digital-choice with face-to-face

|  | **Digital only vs face-to-face**  **Matching rate: 95.0%** | | **Digital choice vs face-to-face**  **Matching rate: 95.9%** | |
| --- | --- | --- | --- | --- |
|  | **Digital only (N=787)** | **Face-to-face (N=2312)** | **Digital only (N=612)** | **Face-to-face (N=1783)** |
| **Sex**  Male, N (%) Female, N (%) | 372 (47.3%)  415 (52.7%) | 1085 (46.9%)  1227 (53.1%) | 289 (47.2%)  323 (52.8%) | 844 (47.3%)  939 (52.7%) |
| **Age at referral**  Mean (SD) Median (IQR) | 61.5 (10.6)  62 (54, 70) | 62.5 (10.3)  63 (56, 70) | 61.1 (10.6)  62 (54, 69) | 62.3 (10.3)  63 (56, 70) |
| **Ethnicity***  White, N (%)  Mixed, N (%)  Asian, N (%)  Black, N (%)  Other, N (%) | 643 (81.7%)  6 (0.76%)  96 (12.2%)  42 (5.34%)  (0.0%) | 1920 (83.0%)  13 (0.56%)  255 (11.0%)  124 (5.36%)  0 (0.0%) | 525 (85.8%)  10 (1.63%)  56 (9.15%)  19 (3.10%)  2 (0.33%) | 1555 (87.2%)  20 (1.12%)  149 (8.36%)  53 (2.97%)  6 (0.34%) |
| **IMD Quintile**  1 (Most deprived)  2  3  4  5 (Least deprived) | 91 (11.6%)  171 (21.7%)  250 (31.8%)  169 (21.5%)  106 (13.5%) | 263 (11.4%)  498 (21.5%)  741 (32.1%)  499 (21.6%)  311 (13.5%) | 145 (23.7%)  79 (12.9%)  87 (14.2%)  139 (22.7%)  162 (26.5%) | 422 (23.7%)  224 (12.6%)  257 (14.4%)  404 (22.7%)  476 (26.7%) |
| **Weight in kg at baseline**  Mean (SD) Median (IQR) | 87.2 (19.0)  85.0 (73.0, 99.0) | 84.5 (18.9)  82.5 (71.2, 95.8) | 86.1 (18.9)  84.5 (72.3, 97.4) | 85.2 (18.8)  83.0 (71.6, 96.2) |
| **BMI at baseline**  Mean (SD) Median (IQR) | 30.9 (6.0)  29.8 (26.6, 34.3) | 30.2 (5.9)  29.3 (26.2, 33.4) | 30.3 (5.88)  29.3 (26.1, 33.2) | 30.4 (6.0)  29.5 (26.3, 33.5) |

* Asian’ comprises those reporting Indian, Pakistani, Bangladeshi, Chinese or ‘other Asian’ ethnicity; ‘Black’ comprises those reporting Caribbean, African or ‘other Black’ ethnicity; ‘Mixed’ comprises people with a Mixed ethnic background and ‘Other’ comprises those reporting any other ethnicity.

**Interaction analyses – digital choice**

Table S5 Complete case interaction analyses assessing differential change in weight from baseline to 6 months between the face-to-face cohort and the digital-choice cohort. Subgroup effects are also shown, where relevant

|  | **Change in weight* (N=30854)** | | | **Change in HbA1c**^†^ **(N=12698)** | | |
| --- | --- | --- | --- | --- | --- | --- |
|  | **B** | **95% CI** | **p-value** | **B** | **95% CI** | **p-value** |
| **Sex**  Male (ref) Female  Interaction | -0.829  -1.298  -0.468 | (-1.397, -0.262)  (-1.862, 0.734)  (-1.019, 0.083) | 0.096 | -0.611  -0.450  0.160 | (-1.434, 0.213)  (-1.270, 0.370)  (-0.470, 0.790) | 0.618 |
| **Age at referral**  Interaction | -0.050 | (-0.074, -0.026) | <0.001 | 0.004 | (-0.024, 0.031) | 0.795 |
| **Ethnicity**^‡^  White (ref)  Mixed  Asian  Black  Other  Interaction (Mixed)  Interaction (Asian)  Interaction (Black)  Interaction (Other) | -1.259  0.995  -0.221  1.749  -0.861  0.263  1.038  3.008  0.398 | (-1.765, -0.752)  (-2.797, 0.806)  (-1.188, 0.747)  (0.104, 3.395)  (-4.647, 2.925)  (-1.521, 2.047)  (0.105, 1.971)  (1.395, 4.621)  (-3.379, 4.175) | 0.772  0.029  <0.001  0.837 | -0.601  -0.108  0.265  0.247  0.503  0.493  0.336  0.849  1.105 | (-1.372, 0.169)  (-2.191, 1.975)  (1.511, 0.981)  (-1.579, 2.073)  (-3.797, 4.804)  (-1.502, 2.489)  (-0.759, 1.432)  (-0.859, 2.556)  (-3.155, 5.365) | 0.643  0.547  0.330  0.611 |
| **IMD**  1 (most deprived) (ref)  2  3  4  5 (least deprived)  Interaction (2)  Interaction (3)  Interaction (4)  Interaction (5) | -0.317  -0.789  -0.568  -1.618  -1.431  0.471  -0.251  -1.301  -1.114 | (-1.057, 0.422)  (-1.660, 0.083)  (-1.439, 0.303)  (-2.311, -0.925)  (-2.094, -0.769)  (-1.423, 0.481)  (-1.237, 0.736)  (-2.139, -0.462)  (-1.953, -0.275) | 0.332  0.618  0.002  0.009 | -0.663  -0.762  -0.035  -0.308  -0.731  -0.098  0.628  0.356  -0.067 | (-1.654, 0.327)  (-1.862, 0.338)  (-1.168, 1.098)  (-1.257, 0.641) (-1.652, 0.191)  (-1.150, 0.953)  (-0.509, 1.765)  (-0.603, 1.314)  (-1.038, 0.904) | 0.854  0.279  0.467  0.892 |
| **Weight at baseline**  Interaction | n/a |  |  | -0.004 | (-0.020, 0.013) | 0.646 |

*All models adjust for age at referral, sex, ethnicity (white/mixed/black/Asian/other), IMD quintile, time since baseline (in months) as fixed effects and CCG nested within STP as random effects
† All models adjust for age at referral, sex, ethnicity (white/mixed/black/Asian/other), IMD quintile, time since baseline (in months) and baseline weight as fixed effects and CCG nested within STP as random effects
‡ Asian’ comprises those reporting Indian, Pakistani, Bangladeshi, Chinese or ‘other Asian’ ethnicity; ‘Black’ comprises those reporting Caribbean, African or ‘other Black’ ethnicity; ‘Mixed’ comprises people with a Mixed ethnic background and ‘Other’ comprises those reporting any other ethnicity.

Older age was associated with greater weight loss in the digital-choice programme in comparison to the face-to-face programme (Table S6).

As for the comparison of face-to-face and digital-only, evidence of a difference across ethnic groups was seen. White individuals lost more weight on the digital programme than the face-to-face programme and Black individuals lost more weight on the face-to-face programme than the digital programme. The interaction effect between White and Black ethnic groups was 3.008 (95% CI: 1.395, 4.621) and the interaction effect between White and Black individuals was 1.038 (95% CI: 0.105, 1.971) suggesting that the difference in weight loss between White and Black individuals and White and Asian individuals was statistically significantly larger in the digital programme than the face-to-face programme. Again, results should be viewed with caution due to the small number of Black and Asian individuals in the digital cohort.

On average, weight loss was greater in the digital-choice group compared to the face-to-face group in all deprivation quintiles: this difference was smaller in magnitude and not statistically significant for quintiles 1, 2 and 3 yet larger in magnitude and statistically significant in quintiles 4 and 5: individuals in quintile 4, on average, lost 1.618kg (95% CI: 0.925, 2.311) more weight on the digital programme compared to the face-to-face programme and individuals in quintile 5, on average, lost 1.431kg (95% CI: 0.769, 2.094) more weight on the digital programme compared to the face-to-face programme. The interaction effects comparing quintiles 4 and 5 with quintile 1 were statistically significant and suggested the difference in weight loss between the least deprived and the most deprived was larger in the digital programme than the face-to-face programme.

**Additional analyses**

***Analyses using regression adjustment to account for confounding***

We re-ran the main analysis using regression adjustment to account for confounding instead of matching. The confounding variables adjusted for were sex, age at referral, deprivation, ethnicity and baseline weight (for the HbA1c analyses only). Again, mixed effects linear regression was used additionally adjusting for CCG nested within STP as random effects. Results are shown in Tables S7 and S8.

Table S6 Regression analyses comparing change in weight and HbA1c from baseline to 6 months and 12 months between the face-to-face cohort and the digital-only cohort using regression adjustment to account for confounding

|  | **N** | **B*** | **95% CI** | **p-value** |
| --- | --- | --- | --- | --- |
| **Weight at 6 months**  Unadjusted  Adjusted^†^ | 33769  31064 | -0.204  -0.136 | (-0.449, 0.041)  (-0.620, 0.347) | 0.102  0.581 |
| **HbA1c at 6 months**  Unadjusted  Adjusted^‡^ | 14843  12843 | 0.330  0.298 | (0.044, 0.616)  (-0.388, 0.983) | 0.024  0.394 |
| **Weight 12 months**  Unadjusted  Adjusted^†^ | 24276  22278 | 0.142  -0.423 | (-0.177, 0.462)  (-0.984, 0.138) | 0.382  0.140 |
| **HbA1c 12 months**  Unadjusted  Adjusted^‡^ | 10361  9016 | 0.715  -0.247 | (0.405, 1.026)  (-1.115, 0.622) | <0.001  0.578 |

*Coefficient quantifies the difference in mean change between the face-to-face and digital cohort, using the face-to-face cohort as the reference group
† Model adjusts for age at referral, sex, ethnicity (white/mixed/black/Asian/other), IMD quintile, time since baseline (in months) as fixed effects and CCG nested within STP as random effects
‡ Model adjusts for age at referral, sex, ethnicity (white/mixed/black/Asian/other), IMD quintile, time since baseline (in months) and baseline weight as fixed effects and CCG nested within STP as random effects

Table S7 Regression analyses comparing change in weight and HbA1c from baseline to 6 months and 12 months between the face-to-face cohort and the digital-choice cohort using regression adjustment to account for confounding

|  | **N** | **B*** | **95% CI** | **p-value** |
| --- | --- | --- | --- | --- |
| **Weight at 6 months**  Unadjusted  Adjusted^†^ | 33574  30854 | -0.943  -1.067 | (-1.214, -0.671)  (-1.566, -0.568) | <0.001  <0.001 |
| **HbA1c at 6 months**  Unadjusted  Adjusted^‡^ | 14728  12698 | -0.195  -0.512 | (-0.499, 0.109)  (-1.247, 0.223) | 0.209  0.172 |
| **Weight 12 months**  Unadjusted  Adjusted^†^ | 24223  22203 | -0.347  -1.001 | (-0.677, -0.017)  (-1.522, -0.497) | 0.039  <0.001 |
| **HbA1c 12 months**  Unadjusted  Adjusted^‡^ | 10182  8826 | 0.625  0.093 | (-0.280, 0.970)  (-0.960, 1.145) | <0.001  0.863 |

*Coefficient quantifies the difference in mean change between the face-to-face and digital cohort, using the face-to-face cohort as the reference group
† Model adjusts for age at referral, sex, ethnicity (white/mixed/black/Asian/other), IMD quintile, time since baseline (in months) as fixed effects and CCG nested within STP as random effects
‡ Model adjusts for age at referral, sex, ethnicity (white/mixed/black/Asian/other), IMD quintile, time since baseline (in months) and baseline weight as fixed effects and CCG nested within STP as random effects

Overall, the results obtained using matching and regression adjustment are very similar and lead to the same conclusions.

At both 6 and 12 months, weight loss in the digital-only and face-to-face cohorts was similar and the upper limits of the 95% confidence intervals for the adjusted mean differences were below the pre-specified non-inferiority limits, demonstrating non-inferiority. Again, the 95% confidence intervals for the adjusted mean differences in HbA1c change were wide and likely do not rule out non-inferiority.

At both 6 and 12 months, weight loss in the digital-choice group was approximately 1kg greater than that in the face-to-face cohort, on average, and this was statistically significant. It follows that non-inferiority was demonstrated. Estimates for the change in HbA1c analyses were also very similar across the two analysis methods. At 6 months, the upper limit of the 95% confidence interval for the difference in reduction in HbA1c was low, suggesting non-inferiority may have been demonstrated for this outcome in this group. However, at 12 months, the upper limit of the 95% confidence interval was above 1mmol/mol suggesting non-inferiority was not demonstrated at this time point.

***Analyses using multiple imputation to account for missing data***

The main analysis was a complete case analysis whereby only observations with complete data were used. The analysis was additionally run using multiple imputation, imputing missing baseline data in both cohorts, as well as missing outcome data in the face-to-face cohort for individuals known to be participating in the programme at the time the measurement was expected.

In the digital cohort, missing baseline information was imputed for individuals who had an observed baseline and 6/12m value for either weight or HbA1c. Regression imputation was used to impute missing values in variables with a small amount of missing data: 6 missing sex values, 1 missing age value and 3 missing IMD quintile values. Multiple imputation by chained equations was used to impute 136 missing ethnicity values.

In the face-to-face cohort, as well as imputing missing baseline information, multiple imputation was to impute missing outcome values for individuals who were still attending the programme at the time of the measurement in the face-to-face cohort.

1. 6m weight analysis. In the complete case adjusted regression analysis, there were 30090 individuals in the face-to-face cohort. Regression imputation was used to impute missing values in variables with a small amount of missing data: 71 missing sex values and 68 missing IMD quintile values (there were no missing age values). Multiple imputation by chained equations was used to impute 2745 ethnicity values, 1612 baseline weight values and 752 6m weight values, resulting in a sample of 34800.
2. 12m weight analysis. In the complete case adjusted regression analysis, there were 21503 individuals in the face-to-face cohort. Regression imputation was used to impute missing values in variables with a small amount of missing data: 71 missing sex values and 68 missing IMD quintile values (there were no missing age values). Multiple imputation by chained equations was used to impute 2021 ethnicity values, 1182 baseline weight values and 619 12m weight values, resulting in a sample of 25068.
3. 6m HbA1c analysis. In the complete case adjusted regression analysis, there were 11936 individuals in the face-to-face cohort. Regression imputation was used to impute missing values in variables with a small amount of missing data: 72 missing sex values and 68 missing IMD quintile values (there were no missing age values). Multiple imputation by chained equations was used to impute 2781 ethnicity values, 15969 baseline HbA1c values, 8902 6m HbA1c values, and 1638 baseline weight values, resulting in a sample of 35407.
4. 12m HbA1c analysis. In the complete case adjusted regression analysis, there were 8188 individuals in the face-to-face cohort. Regression imputation was used to impute missing values in variables with a small amount of missing data: 35 missing sex values and 42 missing IMD quintile values (there were no missing age values). Multiple imputation by chained equations was used to impute 1520 ethnicity values, 7536 baseline HbA1c values, 2936 12m HbA1c values, and 923 baseline weight values, resulting in a sample of 18574.

Table S9 and S10 show the results of the regression analyses comparing change in weight and HbA1c from baseline to 6 and 12 months between the face-to-face cohort and the digital-only and digital-choice cohorts respectively. Differences in sample size between the unadjusted and adjusted analyses are due to missing CCG or STP values as these were not imputed.

Table S8 Regression analyses using multiple imputation comparing change in weight and HbA1c from baseline to 6 months and 12 months between the face-to-face cohort and the digital-only cohort

|  | **N** | **B*** | **95% CI** | **p-value** |
| --- | --- | --- | --- | --- |
| **Weight at 6 months**  Unadjusted  Adjusted^†^ | 35825  35802 | -0.200  -0.181 | (-0.445, 0.044)  (-0.642, 0.280) | 0.108  0.442 |
| **HbA1c at 6 months**  Unadjusted  Adjusted^‡^ | 36362  36341 | 0.226  0.253 | (-0.055, 0.508)  (-0.258, 0.764) | 0.115  0.332 |
| **Weight 12 months**  Unadjusted  Adjusted^†^ | 25886  25862 | 0.142  -0.243 | (-0.177, 0.460)  (-0.785, 0.299) | 0.383  0.380 |
| **HbA1c 12 months**  Unadjusted  Adjusted^‡^ | 19453  19427 | 0.550  -0.169 | (0.239, 0.861)  (-0.785, 0.447) | 0.001  0.590 |

*Coefficient quantifies the difference in mean change between the face-to-face and digital cohort, using the face-to-face cohort as the reference group
† Model adjusts for age at referral, sex, ethnicity (white/mixed/black/Asian/other), IMD quintile, time since baseline (in months) as fixed effects and CCG nested within STP as random effects
‡ Model adjusts for age at referral, sex, ethnicity (white/mixed/black/Asian/other), IMD quintile, time since baseline (in months) and baseline weight as fixed effects and CCG nested within STP as random effects

Table S9 Regression analyses using multiple imputation comparing change in weight and HbA1c from baseline to 6 months and 12 months between the face-to-face cohort and the digital-choice cohort

|  | **N** | **B*** | **95% CI** | **p-value** |
| --- | --- | --- | --- | --- |
| **Weight at 6 months**  Unadjusted  Adjusted^†^ | 35630  35625 | -0.939  -0.935 | (-1.209, -0.668)  (-1.403, -0.468) | <0.001  <0.001 |
| **HbA1c at 6 months**  Unadjusted  Adjusted^‡^ | 36247  36237 | -0.299  -0.287 | (-0.598, 0.001)  (-0.806, 0.232) | 0.051  0.279 |
| **Weight 12 months**  Unadjusted  Adjusted^†^ | 25833  25828 | -0.348  -0.720 | (-0.676, -0.019)  (-1.214, -0.225) | 0.038  0.004 |
| **HbA1c 12 months**  Unadjusted  Adjusted^‡^ | 19274  19267 | 0.459  0.276 | (0.113, 0.805)  (-0.347, 0.900) | 0.009  0.385 |

*Coefficient quantifies the difference in mean change between the face-to-face and digital cohort, using the face-to-face cohort as the reference group
† Model adjusts for age at referral, sex, ethnicity (white/mixed/black/Asian/other), IMD quintile, time since baseline (in months) as fixed effects and CCG nested within STP as random effects
‡ Model adjusts for age at referral, sex, ethnicity (white/mixed/black/Asian/other), IMD quintile, time since baseline (in months) and baseline weight as fixed effects and CCG nested within STP as random effects

In comparison to the results from the complete case analysis, the point estimates are similar but the results are more precise as the sample size has been substantially increased with the multiple imputation. However, the confidence intervals around the estimates regarding change in HbA1c are still wide, so overall conclusions are the same.

Tables S11 and S12 show the results of the interaction analyses comparing the difference in change in weight and HbA1c across the baseline variables between the face-to-face and digital-only and digital-choice cohorts respectively.

Table S10 Interaction analyses using multiple imputation assessing differential change in weight from baseline to 6 months between the face-to-face cohort and the digital-only cohort. Subgroup effects are shown, where relevant

|  | **Change in weight* (N=35802)** | | | **Change in HbA1c**^†^ **(N=36341)** | | |
| --- | --- | --- | --- | --- | --- | --- |
|  | **B** | **95% CI** | **p-value** | **B** | **95% CI** | **p-value** |
| **Sex**  Male (ref) Female  Interaction | 0.394  -0.634  -1.029 | (-0.110, 0.899) (-1.108, -0.161)  (-1.514, -0.543) | <0.001 | 0.296  0.207  -0.089 | (-0.299, 0.892)  (-0.369, 0.784)  (-0.663, 0.485) | 0.761 |
| **Age at referral**  Interaction | 0.006 | (-0.016, 0.027) | 0.622 | 0.013 | (-0.013, 0.039) | 0.317 |
| **Ethnicity**^‡^  White (ref)  Mixed  Asian  Black  Other  Interaction (Mixed)  Interaction (Asian)  Interaction (Black)  Interaction (Other) | -0.464  -0.190  0.534  1.080  -2.820  0.275  0.998  1.544  -2.356 | (-0.931, 0.002)  (-2.327, 1.948)  (-0.285, 1.353) (-0.081, 2.241) (-7.982, 2.342)  (-1.867, 2.416)  (0.122, 1.874)  (0.335, 2.753)  (-7.523, 2.812) | 0.801  0.026  0.012  0.372 | 0.278  0.471  -0.056  0.580  -2.079  0.193  -0.333  0.302  -2.357 | (-0.281, 0.836)  (-1.716, 2.658)  (-1.001, 0.890)  (-0.624, 1.783)  (-8.106, 3.947)  (-1.998, 2.385)  (-1.307, 0.641)  (-0.935, 1.539)  (-8.389, 3.675) | 0.863  0.502  0.633  0.444 |
| **IMD**  1 (most deprived) (ref)  2  3  4  5 (least deprived)  Interaction (2)  Interaction (3)  Interaction (4)  Interaction (5) | -0.686  0.417  -0.246  -0.530  -0.001  1.102  0.439  0.155  0.685 | (-1.422, 0.051)  (-0.200, 1.034)  (-0.831, 0.339)  (-1.188, 0.127)  (-0.771, 0.769)  (0.270, 1.934)  (-0.388, 1.267)  (-0.725, 1.035)  (-0.291, 1.661) | 0.009  0.298  0.729  0.169 | 0.506  0.474  0.299  -0.101  -0.108  -0.032  -0.206  -0.607  -0.614 | (-0.358, 1.369)  (-0.262, 1.209)  (-0.404, 1.002)  (-0.884, 0.682)  (-1.012, 0.795)  (-1.010, 0.945)  (-1.187, 0.773)  (-1.644, 0.429) (-1.752, 0.524) | 0.949  0.679  0.251  0.290 |
| **Weight at baseline**  Interaction | n/a |  |  | 0.007 | (-0.008, 0.022) | 0.372 |

*All models adjust for age at referral, sex, ethnicity (white/mixed/black/Asian/other), IMD quintile, time since baseline (in months) as fixed effects and CCG nested within STP as random effects
† All models adjust for age at referral, sex, ethnicity (white/mixed/black/Asian/other), IMD quintile, time since baseline (in months) and baseline weight as fixed effects and CCG nested within STP as random effects
‡ Asian’ comprises those reporting Indian, Pakistani, Bangladeshi, Chinese or ‘other Asian’ ethnicity; ‘Black’ comprises those reporting Caribbean, African or ‘other Black’ ethnicity; ‘Mixed’ comprises people with a Mixed ethnic background and ‘Other’ comprises those reporting any other ethnicity.

Table S11 Interaction analyses using multiple imputation assessing differential change in weight from baseline to 6 months between the face-to-face cohort and the digital-choice cohort. Subgroup effects are shown, where relevant

|  | **Change in weight* (N=35802)** | | | **Change in HbA1c**^†^ **(N=36237)** | | |
| --- | --- | --- | --- | --- | --- | --- |
|  | **B** | **95% CI** | **p-value** | **B** | **95% CI** | **p-value** |
| **Sex**  Male (ref) Female  Interaction | -0.719  -1.105  -0.386 | (-1.250, -0.189) (-1.637, -0.573)  (-0.914, 0.143) | 0.153 | -0.398  -0.180  0.218 | (-1.004, 0.208)  (-0.787, 0.428)  (-0.385, 0.822) | 0.478 |
| **Age at referral**  Interaction | -0.051 | (-0.074, -0.027) | <0.001 | -0.002 | (-0.028, 0.024) | 0.885 |
| **Ethnicity**^‡^  White (ref)  Mixed  Asian  Black  Other  Interaction (Mixed)  Interaction (Asian)  Interaction (Black)  Interaction (Other) | -1.124  -0.858  -0.009  1.608  -0.835  0.266  1.115  2.732  0.289 | (-1.601, -0.648)  (-2.613, 0.898) (-0.929, 0.910) (-0.183, 3.340)  (-4.143, 2.473)  (-1.484, 2.017)  (0.217, 2.012) (0.946, 4.519)  (-3.020, 3.598) | 0.765  0.015  0.003  0.864 | -0.372  0.143  -0.072  0.494  1.177  0.515  0.301  0.867  1.549 | (-0.916, 0.171)  (-1.773, 2.060)  (-1.127, 0.984)  (-1.126, 2.115)  (-2.619, 4.973)  (-1.378, 2.408)  (-0.723, 1.324)  (-0.715, 2.448)  (-2.238, 5.337) | 0.593  0.565  0.283  0.423 |
| **IMD**  1 (most deprived) (ref)  2  3  4  5 (least deprived)  Interaction (2)  Interaction (3)  Interaction (4)  Interaction (5) | -0.195  -0.692  -0.267  -1.500  -1.379  -0.497  -0.071  -1.305  -1.184 | (-0.879, 0.488)  (-1.501, 0.116)  (-1.086, 0.553)  (-2.167, -0.833)  (-2.018, 0.740)  (-1.385, 0.390)  (-1.000, 0.857) (-2.109, -0.500) (-1.986, -0.382) | 0.272  0.880  0.001  0.004 | -0.191  -0.624  0.309  -0.093  -0.639  -0.433  0.500  0.098  -0.448 | (-0.964, 0.582)  (-1.520, 0.272)  (-0.642, 1.259)  (-0.873, 0.687)  (-1.381, 0.103)  (-1.407, 0.541)  (-0.563, 1.562)  (-0.816, 1.012)  (-1.355, 0.459) | 0.384  0.357  0.833  0.333 |
| **Weight at baseline**  Interaction | n/a |  |  | -0.005 | (-0.021, 0.011) | 0.551 |

*All models adjust for age at referral, sex, ethnicity (white/mixed/black/Asian/other), IMD quintile, time since baseline (in months) as fixed effects and CCG nested within STP as random effects
† All models adjust for age at referral, sex, ethnicity (white/mixed/black/Asian/other), IMD quintile, time since baseline (in months) and baseline weight as fixed effects and CCG nested within STP as random effects
‡ Asian’ comprises those reporting Indian, Pakistani, Bangladeshi, Chinese or ‘other Asian’ ethnicity; ‘Black’ comprises those reporting Caribbean, African or ‘other Black’ ethnicity; ‘Mixed’ comprises people with a Mixed ethnic background and ‘Other’ comprises those reporting any other ethnicity.

All interactions effects were very similar to those from the complete case analysis and the conclusions are the same.

***Analyses comparing change from baseline to completion***

A proportion of participants in the face-to-face cohort did not have a weight or HbA1c measure within 8-14 months after baseline because they had either not yet completed the programme or had completed it early. A sensitivity analysis was run comparing change from baseline to completion weight and HbA1c in the face-to-face cohort with change from baseline to 12 month measures in the digital cohort, where all final outcomes were included regardless of when they were measured. The completion HbA1c in the face-to-face cohort was the value measured by the provider on completion of the programme. The completion weight measure was defined as that recorded closest to and within 31 days of the final HbA1c measure where available, or otherwise that recorded at the final session where this was attended, or otherwise classed as missing. In the digital cohort, completion measures were the 12 month measures used in the main analysis, but here there is no restriction on their timing, i.e. they did not have to be measured between 8-14 months after registration.

Table S13 shows the raw changes in weight and HbA1c from baseline to completion in the face-to-face cohort and the two digital cohorts. Change in weight was similar in the face-to-face cohort and the digital-choice cohort, and slightly lower in the digital-only cohort. Change in HbA1c was largest in the face-to-face cohort. Changes were similar to those at 12 months.

Table S12 Summary of weight and HbA1c outcome measures of participants in the face-to-face cohort and digital-only cohort

|  | **Face-to-face** | **Digital only** | **Digital choice** |
| --- | --- | --- | --- |
| **Weight at completion**  N Baseline; Mean (SD)  Completion; Mean (SD) Change; Mean (95% CI) | 21615  82.56 (17.84)  79.33 (17.50)  -3.23 (-3.29, -3.17) | 891  87.30 (18.97)  84.54 (18.85)  -2.76 (-3.17, -2.36) | 833  86.97 (19.61)  83.54 (19.69)  -3.43 (-3.89, -2.97) |
| **HbA1c at completion**  N Baseline; Mean (SD) Completion; Mean (SD) Change; Mean (95% CI) | 12555  41.02 (3.86)  38.74 (3.94)  -2.28 (-2.36, -2.20) | 951  43.92 (1.87)  42.38 (3.27)  -1.55 (-1.74, -1.35) | 783  43.13 (2.44)  41.40 (3.46)  -1.74 (-1.96, -1.51) |

Tables S14 and S15 show the output from the linear mixed model analyses comparing change in weight and HbA1c from baseline to completion between the face-to-face and digital-only and digital-choice cohorts respectively. Overall, changes from baseline to completion were similar to changes from baseline to 12 months.

Table S13 Regression analyses comparing change in weight and HbA1c from baseline to completion between the face-to-face cohort and the digital-only cohort

|  | **N** | **B*** | **95% CI** | **p-value** |
| --- | --- | --- | --- | --- |
| **Weight Completion**  Unadjusted  Adjusted^†^ | 22506  20833 | 0.467  -0.106 | (0.158, 0.775)  (-0.696, 0.424) | 0.003  0.696 |
| **HbA1c Completion**  Unadjusted  Adjusted^‡^ | 13506  11962 | 0.735  -0.233 | (0.451, 1.020)  (-1.038, 0.571) | <0.001  0.570 |

*Coefficient quantifies the difference in mean change between the face-to-face and digital cohort, using the face-to-face cohort as the reference group
† Model adjusts for age at referral, sex, ethnicity (white/mixed/black/Asian/other), IMD quintile, time since baseline (in months) as fixed effects and CCG nested within STP as random effects
‡ Model adjusts for age at referral, sex, ethnicity (white/mixed/black/Asian/other), IMD quintile, time since baseline (in months) and baseline weight as fixed effects and CCG nested within STP as random effects

Table S14 Regression analyses comparing change in weight and HbA1c from baseline to completion between the face-to-face cohort and the digital-choice cohort

|  | **N** | **B*** | **95% CI** | **p-value** |
| --- | --- | --- | --- | --- |
| **Weight Completion**  Unadjusted  Adjusted^†^ | 22448  20745 | -0.202  -0.717 | (-0.521, 0.116)  (-1.199, -0.236) | 0.213  0.004 |
| **HbA1c Completion**  Unadjusted  Adjusted^‡^ | 13338  11764 | 0.546  -0.053 | (0.235, 0.857)  (-1.042, 0.935) | 0.001  0.916 |

*Coefficient quantifies the difference in mean change between the face-to-face and digital cohort, using the face-to-face cohort as the reference group
† Model adjusts for age at referral, sex, ethnicity (white/mixed/black/Asian/other), IMD quintile, time since baseline (in months) as fixed effects and CCG nested within STP as random effects
‡ Model adjusts for age at referral, sex, ethnicity (white/mixed/black/Asian/other), IMD quintile, time since baseline (in months) and baseline weight as fixed effects and CCG nested within STP as random effects

***Analyses changing the time of the baseline measurement in the face-to-face cohort***

The main analysis used the first intervention session as baseline in the face-to-face cohort as this is when participants first received programme content. A sensitivity analysis was run redefining baseline in face-to-face group as the initial assessment as it is plausible that some individuals may have started to make lifestyle changes after this first interaction, prior to the first session attended. Registration was kept as the baseline time point in the digital cohort.

Table S16 shows the raw changes in weight and HbA1c from baseline to 6 and 12 months in the face-to-face cohort and the two digital cohorts. In both the face-to-face and digital-choice cohorts, a reduction in weight and HbA1c, on average, was observed at both 6 and 12 months. In the face-to-face cohort, change from initial assessment to 6 and 12 months in both weight and HbA1c was similar to change from first intervention session shown in the main analysis.

Table S15 Summary of weight and HbA1c outcome measures of participants in the face-to-face cohort and digital-only cohort where initial assessment was the baseline time point in the face-to-face cohort

|  | **Face-to-face** | **Digital only** | **Digital choice** |
| --- | --- | --- | --- |
| **Weight at 6 months**  N Baseline; Mean (SD)  6m; Mean (SD) Change; Mean (95% CI) | 32476  83.52 (18.28)  81.14 (18.01)  -2.38 (-2.42, -2.34) | 1025  87.43 (19.15)  84.37 (18.65)  -3.05 (-3.38, -2.73) | 830  86.98 (19.04)  83.18 (18.85)  -3.79 (-4.16, -3.43) |
| **HbA1c at 6 months**  N Baseline; Mean (SD) 6m; Mean (SD) Change; Mean (95% CI) | 10377  40.59 (3.87)  38.72 (3.90)  -1.87 (-1.95, -1.79) | 955  43.90 (2.04)  42.25 (3.19)  -1.65 (-1.83, -1.47) | 840  43.16 (2.33)  40.98 (3.33)  -2.18 (-2.38, -1.97) |
| **Weight at 12 months**  N Baseline; Mean (SD)  12m; Mean (SD) Change; Mean (95% CI) | 24799  83.05 (17.99)  80.21 (17.77)  -2.85 (-2.90, -2.80) | 818  87.42 (19.07)  84.52 (18.89)  -2.90 (-3.31, -2.48) | 765  86.92 (19.49)  83.53 (19.50)  -3.39 (-3.86, -2.91) |
| **HbA1c at 12 months**  N Baseline; Mean (SD) 12m; Mean (SD) Change; Mean (95% CI) | 8293  40.74 (3.83)  38.59 (3.87)  -2.15 (-2.24, -2.06) | 879  43.94 (1.82)  42.34 (3.28)  -1.60 (-1.80, -1.40) | 700  43.06 (2.51)  41.37 (3.50)  -1.69 (-1.93, -1.45) |

Tables S17 and S18 show the output from the linear mixed model analyses comparing change in weight and HbA1c from baseline to 6 and 12 months between the face-to-face and digital-only and digital-choice cohorts respectively where initial assessment was used as the baseline time point in the face-to-face cohort. Overall, some point estimates are different to that in the main analysis but changing the timing of baseline in the face-to-face cohort has not impacted the overall conclusions.

Table S16 Regression analyses comparing change in weight and HbA1c from baseline to 6 months and 12 months between the face-to-face cohort and the digital-only cohort where initial assessment was the baseline time point in the face-to-face cohort

|  | **N** | **B*** | **95% CI** | **p-value** |
| --- | --- | --- | --- | --- |
| **Weight at 6 months**  Unadjusted  Adjusted^†^ | 33501  30859 | -0.434  -0.381 | (-0.689, -0.178)  (-0.885, 0.122) | 0.001  0.138 |
| **HbA1c at 6 months**  Unadjusted  Adjusted^‡^ | 11332  10291 | 0.251  0.469 | (-0.030, 0.532)  (-0.221, 1.158) | 0.081  0.183 |
| **Weight 12 months**  Unadjusted  Adjusted^†^ | 25617  23610 | 0.175  -0.241 | (-0.158, 0.509)  (-0.820, 0.337) | 0.303  0.414 |
| **HbA1c 12 months**  Unadjusted  Adjusted^‡^ | 9172  8318 | 0.647  -0.219 | (0.347, 0.947)  (-0.977, 0.540) | <0.001  0.572 |

*Coefficient quantifies the difference in mean change between the face-to-face and digital cohort, using the face-to-face cohort as the reference group
† Model adjusts for age at referral, sex, ethnicity (white/mixed/black/Asian/other), IMD quintile, time since baseline (in months) as fixed effects and CCG nested within STP as random effects
‡ Model adjusts for age at referral, sex, ethnicity (white/mixed/black/Asian/other), IMD quintile, time since baseline (in months) and baseline weight as fixed effects and CCG nested within STP as random effects

Table S17 Regression analyses comparing change in weight and HbA1c from baseline to 6 months and 12 months between the face-to-face cohort and the digital-choice cohort where initial assessment was the baseline time point in the face-to-face cohort

|  | **N** | **B**^‡^ | **95% CI** | **p-value** |
| --- | --- | --- | --- | --- |
| **Weight at 6 months**  Unadjusted  Adjusted^†^ | 33306  30649 | -1.172  -1.373 | (-1.456, -0.889)  (-1.881, -0.866) | <0.001  <0.001 |
| **HbA1c at 6 months**  Unadjusted  Adjusted^‡^ | 11217  10146 | -0.275  -0.580 | (-0.573, 0.024)  (-1.291, 0.131) | 0.071  0.110 |
| **Weight 12 months**  Unadjusted  Adjusted^†^ | 25564  23535 | -0.314  -0.701 | (-0.658, 0.031)  (-1.213, -0.171) | 0.074  0.010 |
| **HbA1c 12 months**  Unadjusted  Adjusted^‡^ | 8993  8128 | 0.556  0.003 | (0.223, 0.889)  (-0.826, 0.831) | 0.001  0.995 |

*Coefficient quantifies the difference in mean change between the face-to-face and digital cohort, using the face-to-face cohort as the reference group
† Model adjusts for age at referral, sex, ethnicity (white/mixed/black/Asian/other), IMD quintile, time since baseline (in months) as fixed effects and CCG nested within STP as random effects
‡ Model adjusts for age at referral, sex, ethnicity (white/mixed/black/Asian/other), IMD quintile, time since baseline (in months) and baseline weight as fixed effects and CCG nested within STP as random effects
